# Health service responses and help-seeking for women experiencing violence during outbreaks in low- and middle-income settings: a scoping review

**DOI:** 10.1101/2025.03.19.25324279

**Authors:** Rose Burns, Manuela Colombini, Neha S. Singh, Janet Seeley

**Author notes:** Corresponding author email contact, 15-17 Tavistock Pl, London WC1H 9SH, UK.

## Abstract

During outbreaks women struggle with access to essential health services, including for violence. Services may be disrupted or deprioritised, or women may avoid clinical settings. We conducted a scoping review to understand how health services for violence against women (VAW) were affected in low- and middle-income (LMIC) settings during recent outbreaks, and women’s help-seeking for violence. We reviewed published academic literature reporting on primary research from LMIC settings during recent outbreaks (Ebola, Zika and COVID-19). Four databases were searched: Medline, Embase, Global Health, and Global Index Medicus. Thirty two papers met the inclusion criteria. Data were extracted using a thematic framework focusing on both the supply and demand for services. Experiences during COVID-19 were overrepresented, with no studies identified from other outbreaks. Research spanned 20 countries including a range of services and populations. In the face of lockdowns and reorientation of the health system towards COVID-19, VAW services were restricted or closed despite being essential. Many settings reported that they moved services online or to telehealth, raising digital access issues and safety concerns for women using services from spaces that might be shared with a violent partner or others. Some in-person programme modalities were also adapted, with community volunteers used, or cash assistance provided to survivors. Help-seeking varied, with greater or fewer numbers of survivors presenting at clinical settings, likely indicating fluctuating restrictions. Women experiencing violence often sought help from informal sources (such as community leaders and family). Survivors’ help-seeking was affected by the economic crisis accompanying COVID-19, including food insecurity and transportation challenges. To prepare for future outbreaks research is needed to identify what services are or are not safe and accessible to deliver online, as well as a understanding a broader range of emerging practices for adapting to social distancing, movement and transport restrictions and economic stress.

## Background

Violence against women (VAW) is a global human rights violation and a public health concern that requires a multisectoral response including the health system [1]. It is linked to multiple adverse physical, mental and sexual and reproductive health (SRH) consequences [1]. The connection between VAW and outbreaks has gained attention, particularly during the COVID-19 pandemic, which was accompanied by a ‘shadow’ pandemic of VAW [2]. During this period alone, 31% of women worldwide experienced intimate partner violence (IPV), with the highest prevalence in developing regions [3]. Recent outbreaks have consistently exacerbated violence due to the accompanying economic stress and lockdowns, which trapped women at home with abusive partners [4,5].

During outbreaks, essential health services for women, including those for maternal, and SRH, are often deprioritised or disrupted. Violence against women services such as psychosocial support, medical and multi-sectoral referrals (e.g. housing) may also be affected. Public health emergencies may further limit access to services due to movement restrictions, reduced availability of commodities, increased pressure on the health workforce, service closures, and social distancing measures affecting in-person service delivery [5,6]. Even outside of outbreaks, health policy and responses to VAW remain a low priority. Whilst 80% of countries have multisectoral VAW policies in place, only 34% include VAW response and prevention as a strategic priority, and just 48% have clinical guidelines for the health sector [7]. The economic and security challenges accompanying crises may further exacerbate barriers to help-seeking [5,6]. Whilst normative efforts (e.g. policy briefs) on how to address VAW have accelerated since the COVID-19 pandemic [8,9], significant gaps remain in understanding what has been implemented in low-and middle-income settings (LMIC) as well as how women seek help [10].

Our study aimed to identify opportunities for strengthening VAW health services during outbreaks in LMICs. We conducted a scoping review of literature on both the supply and demand for health services for VAW during the last decade of outbreaks of emerging/re-emerging pathogens such as Ebola, Zika and COVID-19. We included articles on healthcare, health policy and community health responses, as well as survivors’ help-seeking behaviours. Our objectives were twofold: i) to examine whether and to what extent health services for VAW have been delivered during recent outbreaks and to identify the challenges and facilitators in service provision (supply side); ii) to understand how women sought help for VAW during outbreaks, including the sources of support they tried to access within both the community and health system (demand side).

## Methods

### Study design

We conducted a scoping review drawing on Arksey and O’Malley’s six stage approach: identifying the research question, identifying relevant studies, study selection, charting the data, collating, summarising and reporting the results [11].

### Search strategy and inclusion and criteria

We searched four databases: Medline, Embase, Global Health, and Global Index Medicus, using search terms related to three concepts: i. health services and health care, ii. violence against women, iii. emergent outbreaks such as COVID-19, Ebola and Zika (see supplementary materials for full search strategy). The search was limited to studies conducted in LMICs between 2014 to May 2024, capturing the timeframe of the largest Ebola outbreak to date (which affected West Africa from 2014-2016) and the public health emergency in the Americas following the Zika outbreak in 2015-2016. We focussed on outbreaks that were potentially linked to VAW and that occurred at a large scale, disrupting the health system. The inclusion/exclusion criteria are summarised in Table 1:

**Table 1:** inclusion/exclusion criteria.

|  | Inclusion criteria | Exclusion criteria |
| --- | --- | --- |
| **Population(s)** | - Women survivors of VAW; members of affected communities; members of organisations providing services for survivors (e.g. health workers)<br>- Study populations in LMICs | - Study populations in high income country settings<br>- Children |
| **Intervention(s)** | - Studies looking at access to, and delivery of, health service interventions for VAW survivors during disease outbreaks (e.g. Ebola, Zika, and COVID-19), including how outbreaks affected health services and survivors' help-seeking.<br>- Healthcare interventions for survivors (e.g. post exposure prophylaxis (PEP), emergency contraception, treatment of injuries, psychological first aid, mental health care, psycho social support, referrals)<br>- Health outreach and awareness programmes related to VAW | - Papers reporting exclusively on non -health VAW interventions, including justice and legal aid; prevention; economic empowerment and livelihood interventions<br>- Papers reporting exclusively on maternal and child health, family planning or HIV/AIDS interventions without GBV/VAW components<br>- Interventions during HIV/AIDS epidemics<br>- Interventions during epidemics that are not emergent (e.g. non-communicable diseases) |
| **Outcome(s)** | - Studies describing health services (as outlined above) to address VAW during outbreak periods | - |
| **Study designs** | - Any study design (qualitative, quantitative) that reports results from primary data collection<br>- Peer-reviewed | - Commentaries, blogs, opinion pieces and other sources not reporting primary data |
| **Date or language criteria** | - Published in English<br>- Studies on contemporary and emergent/re-emergent pathogen outbreaks that have occurred at a large scale in LMICs (i.e. Ebola outbreaks, Zika, COVID-19 )<br>- Studies published from 2014 (timeframe of the West African Ebola outbreak) | - Studies published prior to 2014 |

### Screening

We initially screened potential sources by title and abstract, and then by full text against the inclusion and exclusion criteria. One author (RB) screened all abstracts and potentially eligible full text articles and extracted the data. Double screening by multiple authors took place at two different stages of the review. Two reviewers (MC and JS) screened 32 papers at the title and abstract stage; and 4 papers at the full text screening stage. Regular discussions between the first author and co-authors ensured consistency in applying inclusion and exclusion criteria.

### Data extraction

Data were extracted based on the inclusion criteria to cover both supply and demand side topics using Excel. Information including author, title, year published, country, setting, study design, and study period were also extracted from each article.

### Synthesis

We present data using a thematic synthesis approach. On the ‘supply’ side, we identified three themes related to health service delivery: i. disruptions to VAW health service delivery during COVID-19 ii. adaptations to VAW health service delivery iii. continuation of challenges delivering VAW services from ‘normal’ times. On the ‘demand’ side, we identified three themes related to survivors’ help-seeking: i. patterns of survivors presenting at health services, ii. survivors’ experiences seeking help with informal sources such as community members and family and iii. economic barriers to accessing health services.

## Results

### Search results

The search yielded a total of 4,859 articles which were imported into Rayyan and screened for duplicates, then 3,363 articles were screened for eligibility of which 3,307 were excluded and 56 identified for full text screening. After full text screening 34 articles met the inclusion criteria and were included in the final synthesis (see figure 1).

**Figure 1:**
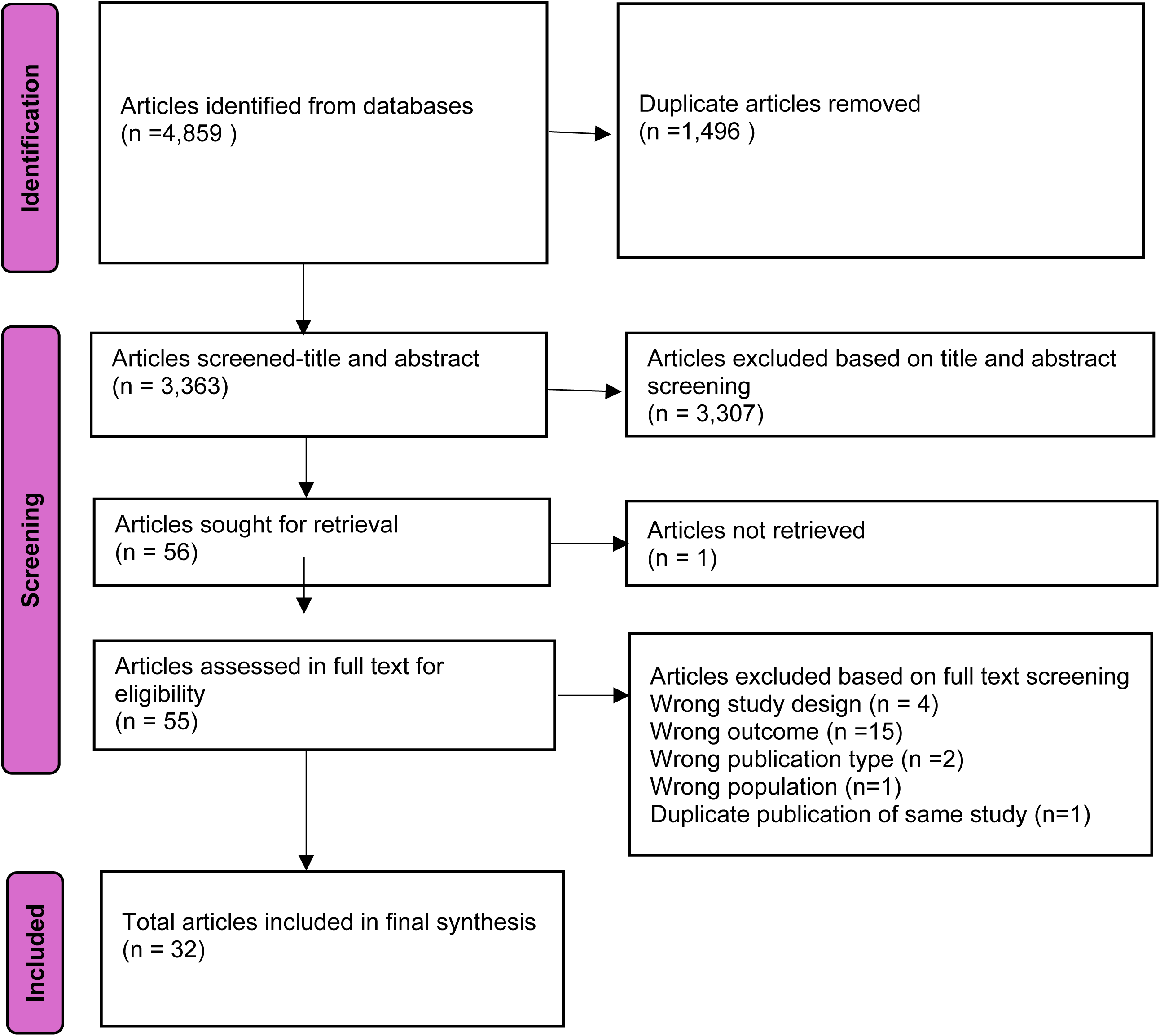
PRISMA diagram.

### Characteristics of studies

All thirty two articles included focussed on experiences during the COVID-19 pandemic and presented both data on VAW help-seeking and on health service delivery for VAW. Studies were published between 2021-2024 with no articles appearing from earlier periods. Studies presented primary research conducted in 20 different countries: India [12–15], Nepal [16,17], Pakistan [18], Bangladesh [18–20], Malaysia [21], Romania [22], the Occupied Palestinian Territory [16], Iraq [23,24], Lebanon [25], Kenya [18,26–32], Mali [16], South Africa [28,33,34], Nigeria [18,28,35–37], Ethiopia [38], Burkina Faso [29], Uganda [27,28], Tanzania [27], Brazil [24,39–41], Guatemala [24,41], and Bolivia [16]. Four articles specifically included or focussed on informal settlements [15,18,24,41], and five on humanitarian settings (e.g. refugee camps or conflict settings) [16,20,23,25,42]. Most studies occurred in a context where VAW, especially IPV, increased (sometimes dramatically) during the pandemic, fuelled by lockdown measures. Many studies simply had data collection take place during the pandemic and it was difficult to disentangle what challenges were specific to the pandemic, compared to pre/post-pandemic. Papers often did not describe locally specific lockdown details. Given the significant variation in these measures across countries and time periods, it was not always clear how different containment measures impacted service delivery or survivors’ help-seeking, especially in retrospective studies.

Articles reported a range of methods, with more than half using qualitative methods. Most studies focussed on violence against cisgender adult women, though some also included child, adolescent or male survivors (with these findings excluded from the review where possible). Many of the studies did not disaggregate the data by age or sex, making it difficult to determine whether the findings were about men, women or children. Studies looked at different types of violence, or different terminology, including VAW, gender-based violence (GBV), sexual and gender-based violence (SGBV), domestic violence, IPV, and sexual violence. Eleven studies explicitly focussed on domestic violence or IPV, which was particularly exacerbated during the pandemic. Studies examined a range of health service delivery settings treating survivors, including both specialised VAW services and other broader healthcare services. Sites included non-governmental organisations (NGO) and government health centres, GBV specific services (e.g. ‘one stop shops’), outpatient departments, community health settings, family health centres, abortion services, counselling and mental health services, and SRH service settings.

Twelve studies presented data on health service delivery (supply topic), nine on help-seeking (demand topic) and eleven had data on both. Most studies provided only minimal findings directly relevant to on our research topic. Instead, they primarily focussed on issues such as increased rates of VAW during the pandemic or the impact of the pandemic on mental health or/and SRH services. While these studies included some incidental data on VAW health service access, only five articles had this as their primary focus [20,24,25,28,38].

We did not conduct a quality assessment of the included studies, in line with scoping review methodology, which focuses on mapping the breadth of the literature rather than assessing the quality of individual study findings.

A description of the 32 articles included in the final review is provided in Table 2:

**Table 2:**
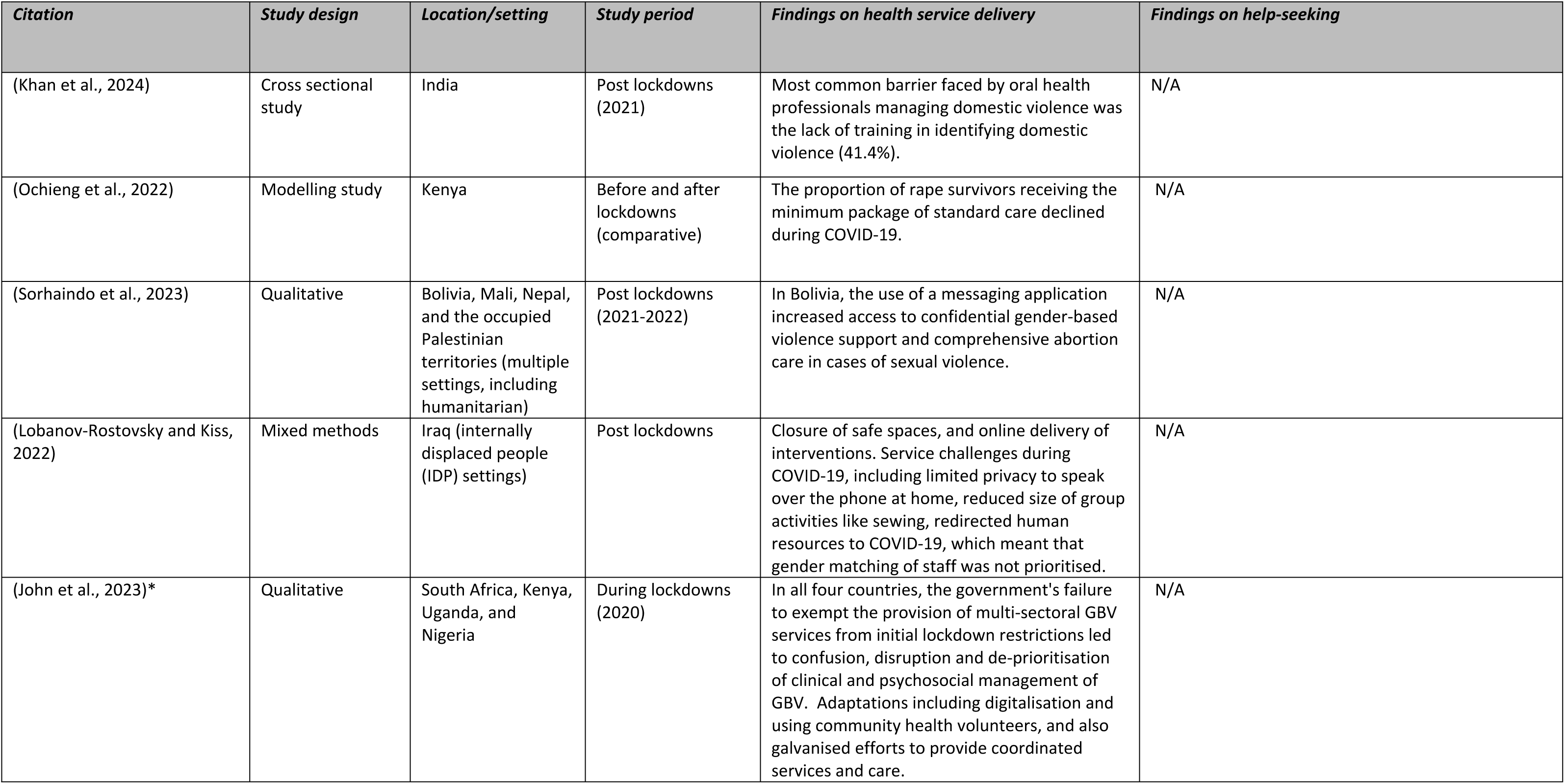
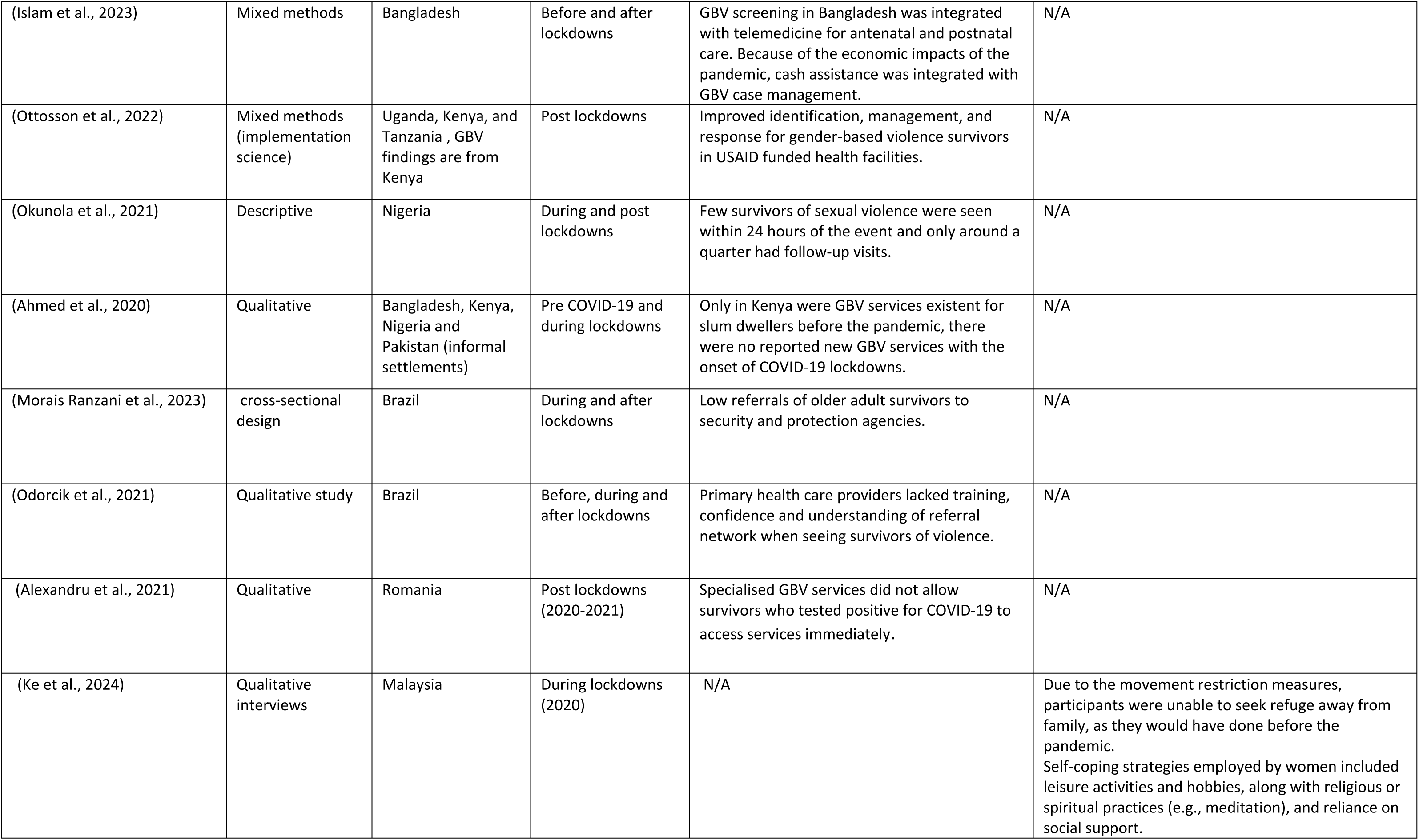
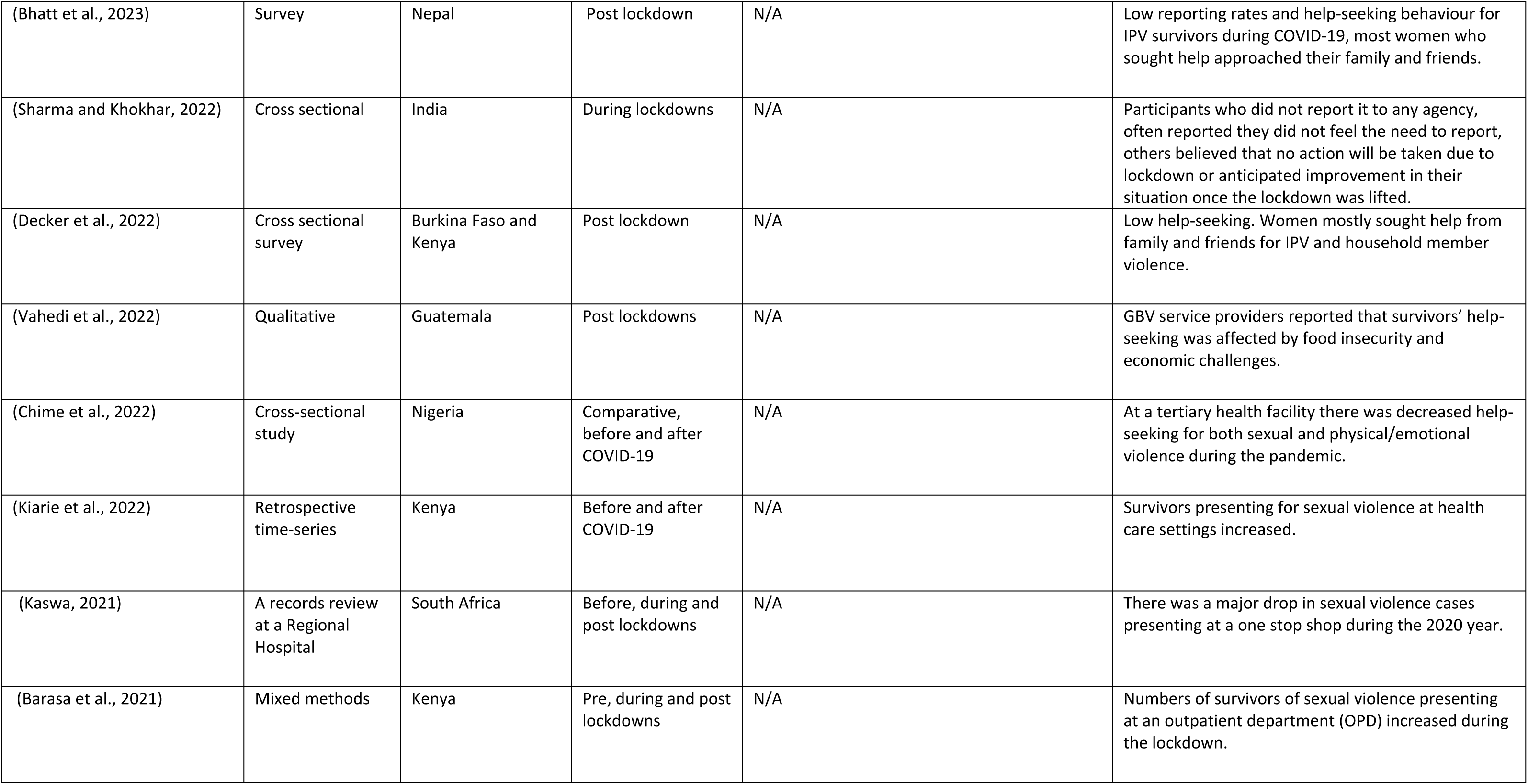
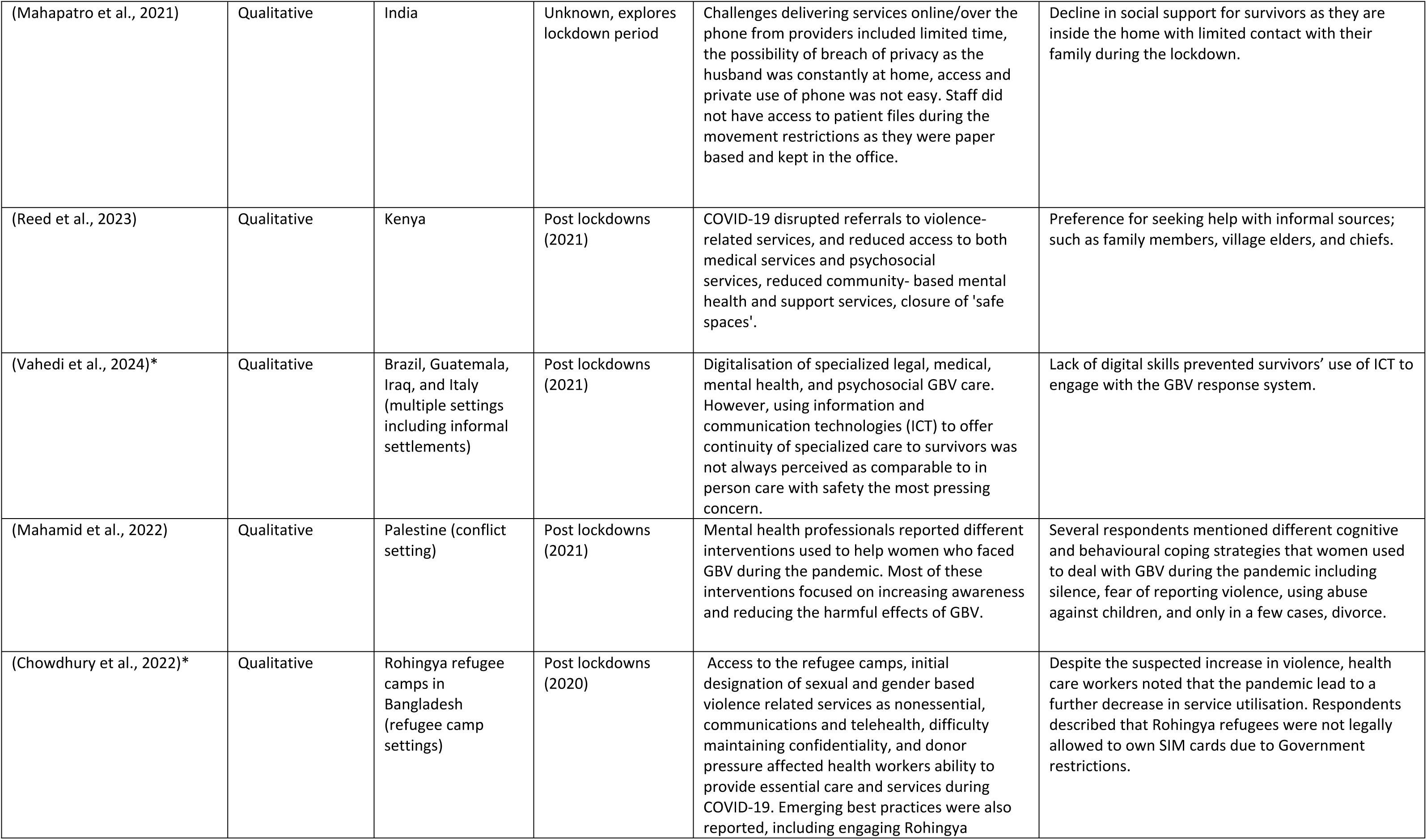
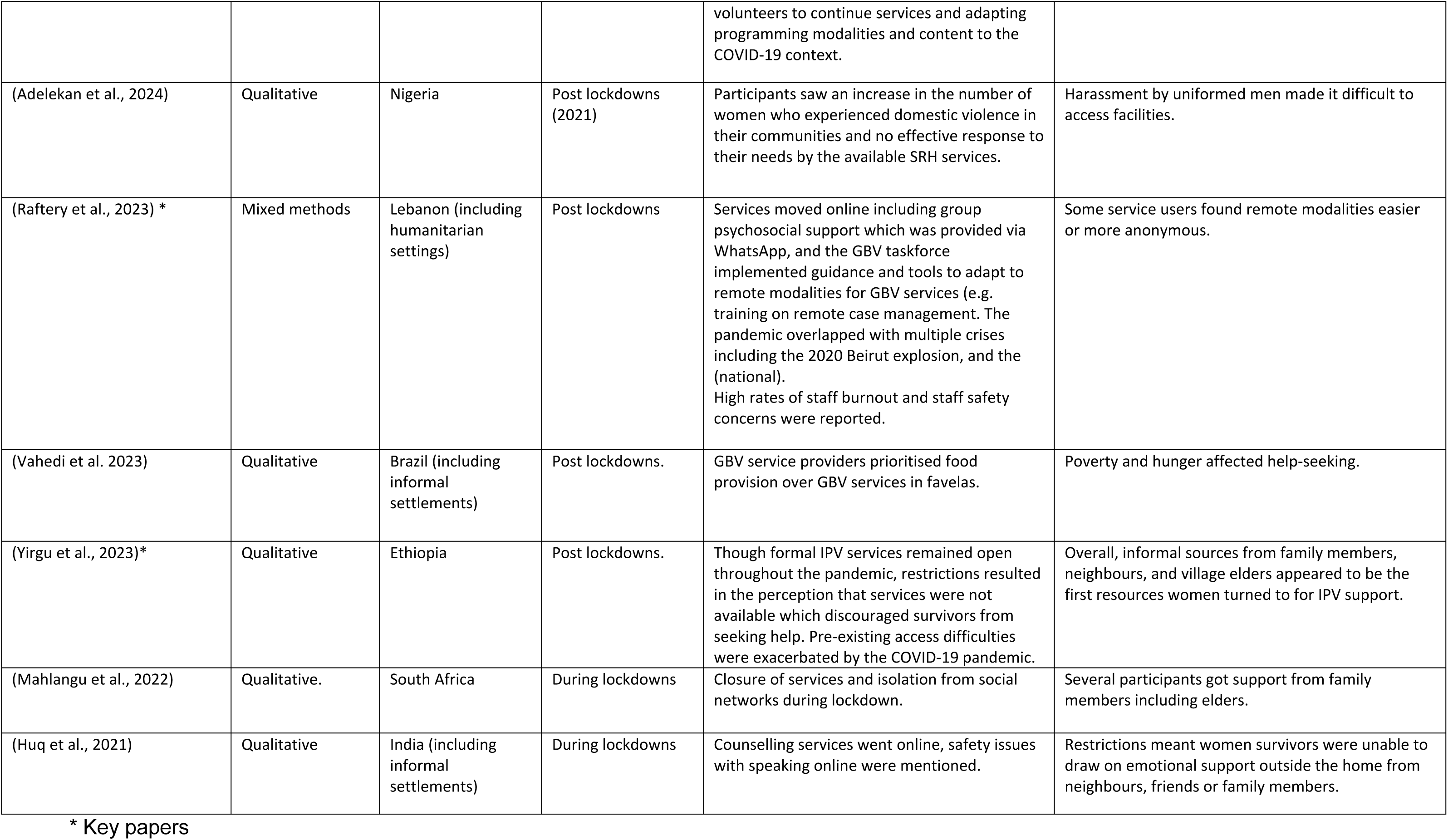
Summary table with study characteristics.

## Results

This section organises the literature thematically into both our supply and demand side topics. First, we present the key findings related to supply themes, focusing on health service delivery for VAW during COVID-19. Then, we present the key findings related to demand themes, highlighting women’s help-seeking during the pandemic.

### 1. Health service delivery for VAW during COVID-19 (supply themes)

#### 1.a. Disruptions to VAW health service delivery during COVID-19

Across settings, in-person services were disrupted or closed down during the COVID-19 pandemic due to the containment measures or the health system being reoriented to the COVID-19 response. Given that some papers lacked details on locally specific lockdown measures, which varied significantly across different weeks and settings, it is difficult to fully understand which factors were more disruptive. In some settings, essential VAW services were severely impacted. In Bangladesh, in Rohingya refugee camps during the pandemic, SGBV services were initially designated as ‘non-essential’ and needed to drastically reduce their staff. Participants noted that this de-prioritisation slowed down service provision for SGBV [20]:

> “The government … [is] giving more priority to both food and health as life-saving. But, that SGBV is also a part, and that it can also be lifesaving, on that we noticed a little indifference on the part of the government….If they would have given priority to the GBV part as a lifesaver, we could have made our work faster.”[GBV service provider]

The containment measures produced multiple additional challenges that affected health workers’ ability to provide essential care and services to Rohingya survivors. This included access to the camps; social distancing, which reduced the number of people inside a service; the types of services possible and how they were delivered (given that three feet of distance needed to be maintained); and donor expectations to continue service delivery despite government restrictions [20]. ‘Safe spaces’ were also closed in non-humanitarian during the pandemic, cutting off access to many VAW related services such as information, social support and ‘mentors’ who could provide counselling [32]:

> “you will find us missing our ways because there is no one to advise us.” [adolescent girl or young woman participant, Kenya]

In other sites, services remained open but were overwhelmed by the demands of the pandemic, leading to inadequate care for VAW survivors. In Lebanon high rates of staff burnout were reported, making it challenging to sustain GBV service provision, staff safety concerns were high with threats and theft reported by most service providers [25]. In Kenya, Uganda, South Africa and Nigeria, GBV service providers reported that government decisions to reassign health workers to the COVID-19 response led to fewer providers available to serve the specific medical needs of survivors, including medical forensic exams [28]. In Kenya, health workers overburdened by COVID-19 cases de-prioritised clinical management of rape cases including medical forensic exams [28]. Survivors who did reach health facilities were sometimes told to come back later for care [28].

Studies reported a decline in the availability and quality of services for survivors. A study using data from the Kenya Health Information System reported a decline in the proportion of rape survivors (all ages and genders) receiving the national minimum package of standard care during the pandemic [26]. For example, the percentage of reported rape survivors receiving recommended HIV post-exposure prophylaxis (PEP) decreased from 61% to 51% [26]. In a study on psychosocial interventions with displaced women survivors in Iraq, human resources were redirected to COVID-19, which meant gender matching of staff was not prioritised, limiting women survivors access to female staff [23]. In other settings like Romania, GBV service providers within NGOs and government settings (in health and non-health) reported that specialised services would not (immediately) accept survivors who tested positive for COVID-19 [22]. In Nigeria adolescents and women of reproductive age reported that with the increases in domestic violence in their communities during the lockdowns there was no effective response to their needs by the available SRH services [37].

There was also an account of VAW interventions delivered successfully despite the pandemic (but potentially with significant resource injection) [27]. A USAID funded project in Kenya focussed on locally lead solutions improved the identification, management, and response for gender-based violence survivors by a monthly average of 642% [27]. This also occurred despite a health worker strike [27].

#### 1.b. Adaptations to VAW health service delivery during COVID-19

##### Moving VAW health service online

Lockdown restrictions on gatherings and social contact curtailed in-person delivery of services across health care settings. In the wake of this, many services, especially counselling, adapted by rapidly moving to delivery online or over mobile phones and this was discussed in nine of the included articles[14–16,19,20,23–25,28]. Safety concerns were the most pressing ethical consequence, and the gender digital divide.

In Kenya and South Africa, participants described providing counselling services remotely to survivors during the lockdowns[28]. In Brazil, Guatemala and Iraq some legal, medical, mental health, and psychosocial care for survivors was digitalised [24]. This included remote legal counsel and group information or counselling sessions for survivors [24]. However the continuity of specialized care to survivors not always perceived as comparable to in person care given that implicit signs of abuse were difficult to spot or document when a survivor is seen online [24]. Several sites mentioned the safety concerns of offering VAW care if perpetrators saw or overheard the reporting or safety planning that happened online or over the phone. In an Indian site, where counselling service moved online, women were hesitant to discuss when their husband could overhear them fearing the repercussions [15], privacy was a similar concern for internally displaced women survivors in Iraq engaged online or over the phone with psychosocial interventions [23]. In Bangladesh many women risked revealing their experiences of SGBV to others when engaging with case workers, volunteers, and other service providers over the phone in public or in their homes [20].

Limited digital access also raised challenges for safe, effective and equitable service provision. GBV screening in Bangladesh was integrated with telemedicine for antenatal and postnatal care in Bangladesh [19]). However both survivors and providers may not have adequate access to internet or telephones (phones may be owned by husband or family members) [19]. Similarly in Brazil, Guatemala and Iraq, challenges included quality information and communication technology (e.g. battery life, headphones), digital skills (e.g. connecting to Wi-Fi, texting etc.) and the cost of data [24].

There were greater challenges in humanitarian settings or for marginalised populations. In Bangladesh, Rohingya refugees, including Rohingya staff/volunteers living in camps, were not legally allowed to own SIM cards due to government telecommunication restrictions implemented pre-pandemic [20]. These restrictions made it difficult for health workers to communicate with patients, and they particularly affected counselling, survivor outreach, and other essential services and referrals. In Nigeria, Kenya, South Africa and Uganda, providers felt that while innovative solutions were attempted, structurally excluded populations such as adolescent girls and young women faced challenges in accessing phones or other virtual technology in a confidential and comfortable way for online counselling [28]. In some cases, digital services could be used for triaging survivors who needed in person services. For instance, in a study on GBV coordination in Lebanon, group psychosocial support was provided via WhatsApp using a mixed approach of chats, voice messages and live calls [25]. However, high-risk cases were prioritised for in-person services [25].

There were some positive reports despite these challenges. In Lebanon some service users preferred remote modalities due to difficulties with transportation and the risk of infection, and there was more anonymity for LGBTIQ + users [25]. Some providers felt women might be more comfortable to disclose violence over the phone [19]. In some settings virtual counselling was managed in groups of four to six women survivors and the ability to work in a group was seen to improve communication and learning between women [24]. In Bolivia a social media campaign focused on educating people about their legal right to medical abortion in cases of sexual violence, and the use of a messaging application confidentially linked survivors with providers for medical abortion [16].

On the other hand, some services operated using paper-based systems and could not transition online. At an Indian NGO, counsellors reported that they could not offer services until the lockdown eased because survivors’ phone numbers and records were kept in the office, which providers were not allowed to access due to movement restrictions [14].

##### In-person VAW health service delivery adaptations

In-person adaptations were also described in terms of guidance, partnerships and the use of new modalities with volunteers and economic interventions used during the pandemic. In Lebanon, the pandemic overlapped with multiple crises including the 2020 Beirut explosion, and the (national) GBV taskforce implemented harmonised guidance and tools for actors at field level to adapt to remote modalities for GBV services (e.g. training on remote case management)[25]. South African providers reported that collaborations were galvanised as smaller local organisations found more ways to connect with larger NGOs and private foundations during the crisis given meetings moved online [28]. In Kenya, given the challenges with digitalising, some organisations experimented with using Community Health Volunteers (CHVs) who were trained to provide psychological first aid in-person during the lockdown and helped refer survivors to health facilities. The approach was successful in serving some harder to reach women, but some providers were concerned that the perceptions that CHVs could not give professional counselling could damage service provision in the future [28]. In Bangladesh when staff could not access camps, engaging Rohingya volunteers, including psycho social support volunteers, was described as an emerging best practice [20]. Volunteers became essential to continuing services by visiting households and referring cases [20].

Programme activities were also pivoted to making face masks (with livelihood components) an activity which included referrals for women experiencing violence [20]. In other sites adaptations included economic interventions for survivors. Interestingly in Brazil, GBV providers used their limited resources for food provision rather than GBV services because of the enormous economic hardship and structural violence that favela communities were facing [41]. Similarly in Bangladesh the United Nations Population Fund (UNFPA) piloted integration of cash assistance within GBV case management services [19]. These new modalities seemed to occur in sites facing extreme economic repercussions from the pandemic.

#### 1.c. Continuation of challenges in delivering VAW health services from ‘normal’ times

In this section we mention studies where challenges encountered by service providers were likely more longstanding, rather than specifically related to the pandemic. These challenges include issues around training and referrals.

In Brazil, primary health care providers lacked training and understanding of referral network when seeing survivors of violence [40]. In a study in India with oral health professionals, whilst over half had screened new patients for visible signs of domestic violence, the most common barrier they encountered in managing survivors was the lack of training in identifying domestic violence (41%) [43]. Similarly, in two Family Health Centres in Brazil, a qualitative study with health professionals found that training was needed to build confidence to properly carry out identification and referral of survivors [40].

Referral by health providers was also a challenge that may have pre-dated the pandemic. A cross-sectional study in Brazil looking at older adult survivors (men and women) shows that for older adults after being seen in hospitals, outpatient services and other public services, there were low referrals to security and protection agencies [39]. In Palestine, counsellors mentioned GBV survivors’ reluctance to engage with mental health services, including perceptions that counselling was for ‘mad’ people, or a preference for medication rather than counselling [42].

In some sites, services for survivors were limited or non-existent regardless of COVID-19. A study with stakeholders in Bangladesh, Kenya, Nigeria and Pakistan showed that only in Kenya were GBV services existent for slum dwellers before the pandemic [18]. In Ethiopia, services, for example from women’s affairs and health facilities, were particularly limited at the local administrative (Kebele) level [38].

### 2. Findings on women’s help-seeking after experiencing violence during COVID-19 (demand themes)

#### 2.a. Patterns of survivor presentation for violence at health services

Studies presented a mixed picture on whether health facilities saw increases or decreases in numbers of VAW survivors presenting for care, perhaps reflective of differing lockdown contexts.

In Kenya, the number of survivors of sexual violence presenting at an outpatient department (OPD) *increased* at a monthly rate of 0.15 cases after March 2020 during the lockdown [44]. In another study in Kenya data was collected from health facilities across the country and compared different time periods including pre-pandemic, two time periods during the pandemic, and a period during which there was a health worker strike [30]. Survivors presenting for sexual violence also *increased* by 8% during the pandemic, even when presentations for other concerns were decreasing (such as OPD visits and HIV testing) [30].

In Nigeria, at a GBV unit at a tertiary health facility, over a three year period there was decreased presentation for both sexual and physical/emotional violence during the pandemic, with a peak pre-pandemic in 2019 and a reduction of cases in 2020 during COVID-19 [36]. Despite reduced help-seeking, the proportion of cases of physical/emotional violence (96%) to the total cases of GBV per year was higher in 2020 during the pandemic compared to 2019 and 2018. This may be connected with the lockdown exacerbating physical violence [36]. In South Africa, at a ‘one stop shop’ for health service delivery for sexual violence there was also a major *decrease* with only about half (451) of the annual average cases of sexual violence presenting in 2020 compared to pre-pandemic [34].

In all four of the studies mentioned above [30,34,36,44], the authors do not specify whether survivors were presenting for IPV or violence perpetrated by non-partners, and the age and gender of survivors is not reported on. Therefore, it is difficult to interpret whether they reflect changes in patterns of violence (e.g. increases/decreases in violence) or they reflect changes in help-seeking. And we are not aware to what extent children were included, who may have faced quite different patterns of violence during stay-at home orders and school closures.

Other than cases seen, there are important indicators of help-seeking such as late attendance and poor follow up, which have implications for time sensitive interventions such as post-exposure prophylaxis for HIV and emergency contraception. In a descriptive study of survivors of sexual assault who received medical care at a ‘one-stop’, free sexual assault clinic in Nigeria from mid-2020 to mid-2021 out of 74 women and girls presenting only seven survivors were seen within 24 hours of the event and around a quarter had follow-up visits [35]. Again this study may be reflective of pre-pandemic trends.

In some cases, such as a cross sectional studies, it was unclear whether the pandemic saw changes in help-seeking given that in many settings survivors rarely seek help from formal services. For example, in one study in Nepal, only 14% of the survivor participants (IPV survivors who were women of reproductive age) reported seeking help [17]. Help-seeking was made difficult by economic stress, fear of separation from children, fear of retaliation from partners, limited family and community support and social isolation, travel restrictions, and higher distress levels during the pandemic [17]. In an Ethiopian site, no pregnant IPV survivors in the study sought help at health services during the height of the COVID-19 pandemic [38]. This was influenced by the fact that, although many formal IPV services remained open throughout the pandemic (police, courts and health centres were mentioned), lockdowns left pregnant IPV survivors and the larger community with the perception that services were not available [38],

#### 2.b. Women’s experiences seeking help with informal sources such as community members and family

Given the challenges in accessing formal health services during COVID-19, and, in some settings reduced help-seeking, many women turned to informal sources like community members or family for support. This trend may be especially true in cases of IPV, influenced by factors that pre-date the pandemic. Family or community support may be sought in the hope of preserving a marriage and avoiding the social consequences of a divorce. Help-seeking from informal sources is described in seven articles [13,15,17,21,29,33,38] including two articles mentioned above (2.a.) where survivors rarely sought help from formal sources. We will first describe the prevalence of help-seeking from community members (which may not be specific to the pandemic period) and then we will explore pandemic-specific disruptions to help-seeking from these community supports as well as survivors’ coping mechanisms.

In Nepal, among IPV survivors (women of reproductive age) who sought help during the pandemic, the most common sources were family (43%), followed by friends (38%), neighbours (10%), and others (10%) [17]. In a cross sectional survey in Burkina Faso and Kenya, over half of women did not seek help for the violence they experienced during the pandemic period [29]. Across violence types, help-seeking concentrated on informal help (32–43%), with the woman’s own family, the husband/partner’s family, or friends, reported as most frequently accessed sources of help [29]. Authors do note this is consistent with pre-pandemic global trends. In South Africa, a qualitative study showed that for IPV during the lockdown couples used (family) elders who gave advice [33]. Similarly, in a study in Ethiopia with pregnant survivors of IPV, family members, neighbours, and village elders appeared to be the first resources women turned to for IPV support [38]. The majority of women described these support systems as useful for encouraging negotiations between partners, as well as offering temporary housing as a way to de-escalate the situation [38].

However, COVID-19 also disrupted help-seeking from informal and community sources of support. Social distancing, stay home orders, and restrictions of on movement also hindered women from receiving support for IPV from neighbours, family, friends, and religious leaders compared to pre-pandemic [38].

> “Before coronavirus I used to tell the mosque Imam and village elders when we have disagreements. But after coronavirus, there were movement restriction, and I could not meet anyone.” [IPV survivor]

In Malaysia women and older adults were unable to seek refuge away from family, as they would have before the pandemic [21] and in India women were less able to draw on emotional support outside the home from neighbours, friends or family members during the pandemic [15]. In some cases survivors simply tried to cope on their own: In India of 7 men and women study participants who faced domestic violence during lockdown, half of the survivors chose to ignore it and around half used yoga/meditation to cope [13]. Participants (men and women) who did not report it to any agency, stated they did not feel the need to report, believed that no action will be taken due to lockdown, or anticipated improvement in their situation once the lockdown was lifted as there would be less interaction with the partner [13].

#### 2.c. Economic barriers to accessing health services

In many settings economic crises accompanied the pandemic and impacted a range of sectors. In two studies in Brazil and Guatemala the pandemic and containment measures meant that survivors, especially marginalised groups such as Black, trans, LGBTQIA+, migrant, sex workers and survivors living in favelas, faced economic stress which affected their help-seeking [41,45]. This included poverty, food insecurity, unemployment, lack of electricity, precarious labour, and weakened transportation infrastructure [41,45]. The authors found that survivors often faced dire food insecurity (as well as water and sanitation needs) meaning that they prioritised basic survival over GBV treatment and care [41]:

> “They are the families in poverty, below the poverty line. They are those families that come, for example, to the place and ask: ‘I just want food. I do not care if I get beaten, if my husband drinks every day, and comes drunk and hits everybody. No, I want food.’”[service provider]

A weakened economic position also made it more difficult to escape violent partners and seek help, and movement restrictions and price surges limited access to public transportation [45]. In African settings a similar picture emerged. In an Ethiopian study distance to services was a barrier during ‘normal’ times, however this worsened during COVID-19 with increased transportation costs making it harder for women attempting to use formal IPV services [38]. In a qualitative study in Nigeria, Kenya, South Africa, and Uganda respondents from the four countries described how restrictions on the use of public transportation constrained or delayed survivors’ access to medical clinics, especially for those living in rural areas [28]:

> “We also have challenges in terms of physical visits because some survivors are not able to access the centre because of restriction in movement.” [Clinical service provider, Nigeria]

Across regions, economic concerns were heightened during the pandemic especially for marginalised groups or those living in rural areas or informal settlements, with food insecurity and transportation challenges affecting survivors’ help-seeking.

## Discussion

To our knowledge, this is the first review to examine health service delivery and help-seeking for VAW during recent outbreaks in LMIC settings. Given that outbreaks exacerbate VAW and disrupt health service access for survivors, evidence synthesis is essential in order to identify opportunities for strengthening service delivery. Although our search focussed on multiple recent outbreaks (Ebola, Zika, COVID-19), all 32 papers included only examined experiences during COVID-19. We found that there was reduced VAW health service delivery during COVID-19 with a variety of services affected by movement restrictions, staff reallocation to the COVID-19 response, lower service quality, being designated as ‘non-essential’, and closure of services such as safe spaces. Some challenges mirrored those that may pre-date the pandemic such as training and referrals. Many articles described how VAW services attempted to adapt during the pandemic. Services, especially counselling, moved online or to telehealth service delivery. However, this presented safety challenges for survivors who did not have private spaces to speak, and for more marginalised groups (populations in humanitarian settings or youth) who may lack digital access. Some services adopted new delivery modalities with volunteers alongside economic interventions used to respond to survivors’ basic needs. Some clinical settings saw more survivors presenting during the pandemic, and some saw fewer, this may have been dependent on the types and timings of lockdowns which discouraged use of the health system. In many cases survivors relied on informal sources of help like their families or community members, and the economic crisis that accompanied the pandemic globally presented challenges to help-seeking with transportation challenges exacerbated by the movement restrictions.

Despite calls over the previous decade to ensure the continuity of essential health service delivery for women during outbreaks, this review shows that across LMIC settings, VAW services were de-prioritised or closed down during COVID-19. Whilst COVID-19 generated political attention to this topic, specifically to the impact of stay at home orders when survivors were locked down with their violent partners, it is not clear whether this will translate into long term investment or policy change. There is a large body of prevalence studies (including reviews) which look at increases in some types of violence during COVID- 19 across various settings [3,46–48]. However, this body of literature does not shine light on what responses were available to survivors during the pandemic and what could be learnt from this for future outbreaks. There is also body of literature looking at the secondary impacts for COVID-19 on the health system more broadly (e.g. routine immunisation, SRH services) [49,50]. Despite significant focus on these two topics, these bodies of research have not been linked with a review: therefore this scoping review provides a timely contribution.

However, many of the papers we included only look at VAW health service delivery as a very small part of their study, therefore further work is needed to build a picture of how LMIC settings might strengthen VAW service delivery during outbreak containment measures.

There is a significant research gap in experiences from the range of other (non-COVID-19) outbreaks that have also shattered heath systems and exacerbated violence. This gap is striking considering that outbreak responses to Ebola (in West Africa and Democratic Republic of the Congo) and Zika (in the Americas) have also been critiqued as gender blind and as having neglected other essential services for women [51–54].

The literature included in our review indicates that many LMIC health systems have significant gaps in terms of resilience and readiness to delivery VAW services during outbreaks like COVID-19. The concept of resilience describes how services continued, or were disrupted, adapted or used different resources in order to deliver services [55]. Most of the adaptations reported during COVID-19 involved remote delivery of counselling services online, and this appeared to be inferior to in-person service delivery as it was fraught with safety challenges and inequitable digital access. In preparation for future outbreaks evidence is needed on digitalisation of VAW services: what can be done safely online and what poses too many safety and access challenges? Gaps remain in our understanding of other adaptations. Whilst a few examples were mentioned there is a need for more research (including that from outbreaks other than COVID-19) to understand emerging best practices for delivering services during social distancing measures, as well as how survivors might manage disruptions to transport, movement restrictions, and pressing economic concerns.

Despite calls to examine outbreaks through a gender lens, we also found, as others have noted [56], that many COVID-19 studies did not present age, gender or sex disaggregated results. This makes it difficult to interpret whether changes in numbers of survivors presenting at health services were reflective of help-seeking patterns or reflective of increases/decreases in specific types of violence. For example, we know that in some settings the risk of violence against children increased, influenced by the closure of schools and increased economic stress inside families [57]. At time, it was unclear whether the health service data in articles in the included articles pertained to children or adults.

### Strengths and limitations

We did not conduct a search of grey literature, which may partly explain why there was no research found on Ebola, Zika or other recent emergent outbreaks. We assume that some research is available in the grey literature such as NGO reports. Similarly, the time period during which we conducted the search was shortly before Mpox was declared a public health emergency of international concern, given there are important gender considerations around Mpox [58] this scoping review missed any literature emerging on this topic.

We also only included articles published in English which might partly explain why we did not identify any literature on non-COVID-19 outbreaks, especially Zika.

### Conclusions

This scoping review has revealed an overwhelming focus on experiences during COVID-19 in the literature. The literature we identified has highlighted that, despite longstanding consensus that life-saving services for survivors of violence need to be provided during outbreaks, such services were deprioritised or closed down in many LMIC settings during the pandemic. Adaptations were mostly around remote service delivery, and these was fraught with safety challenges. To strengthen VAW service delivery during future outbreaks, further work is needed to determine which services can be ethically delivered remotely and online. Further research is also needed to explore a broader range of good practices, including beyond the COVID-19 pandemic, that might have emerged in response to movement restrictions, economic pressure and social distancing in health facilities as well as with women’s help-seeking.

## Data Availability

All relevant data are within the manuscript.

## Acknowledgments

Thank you to Kathleen Perris at the London School of Hygiene and Tropical Medicine library for her support in developing the search strategy.

## CRediT authorship contribution statement

**Rose Burns:** conceptualisation, methodology, formal analysis, investigation, data curation, writing original draft, writing review and editing, visualisation, supervision, project administration. **Manuela Colombini:** conceptualisation, methodology, formal analysis, investigation, writing review and editing, supervision. **Neha Singh:** conceptualisation, methodology, supervision. **Janet Seeley:** conceptualisation, methodology, formal analysis, investigation, writing review and editing, supervision.

All authors reviewed the final manuscript.

## Supplementary materials

**Table 3:**
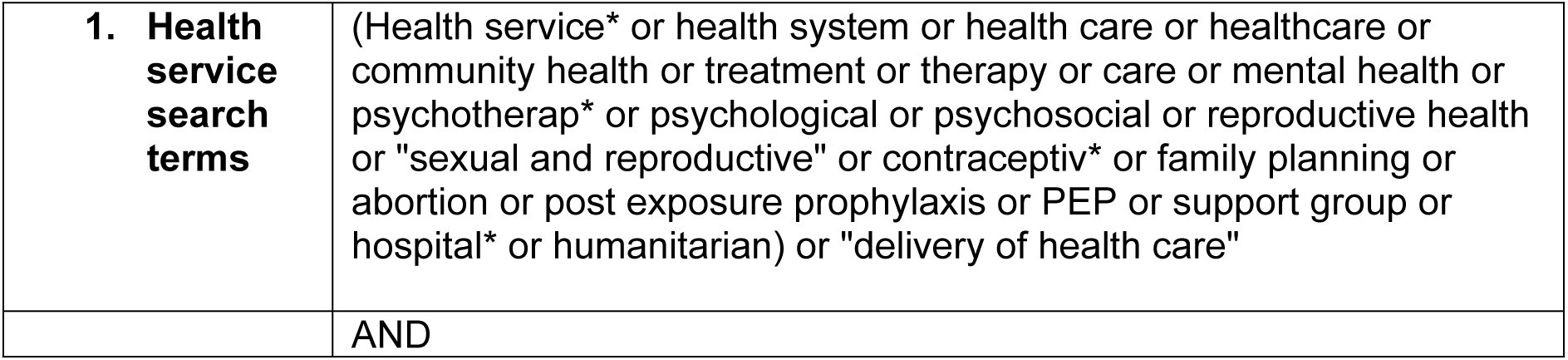
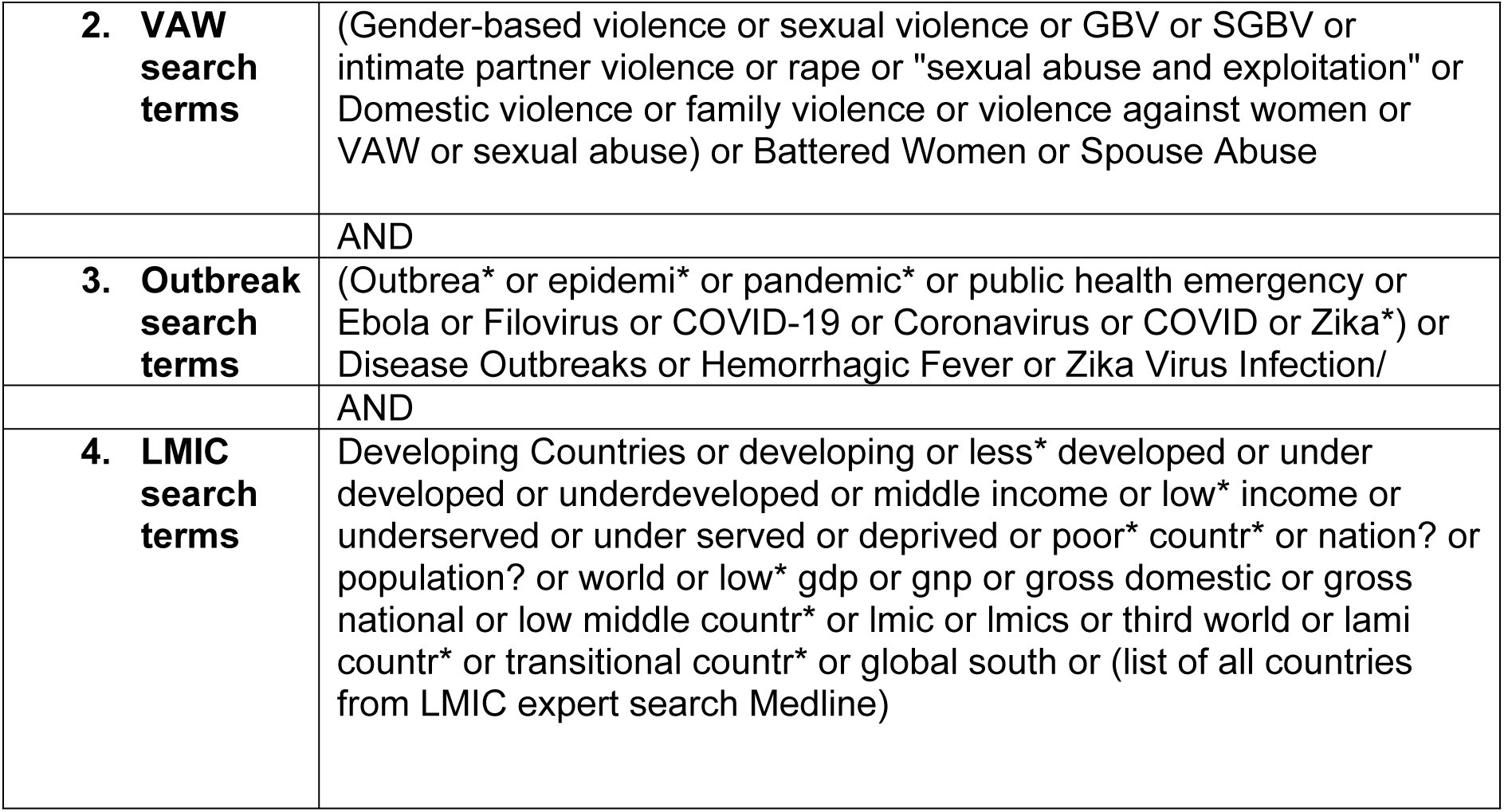
search strategy Medline.

**Table 4:**
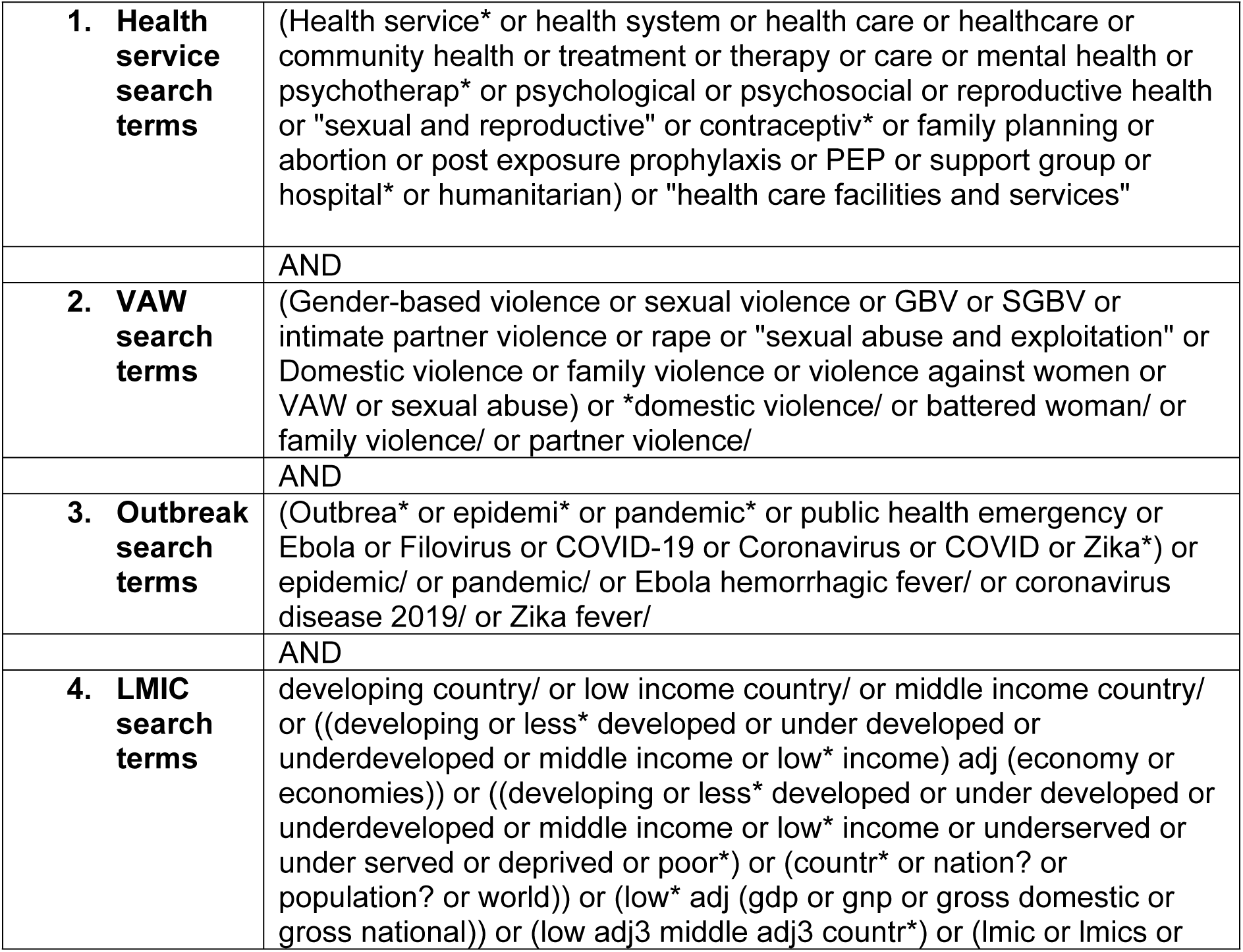
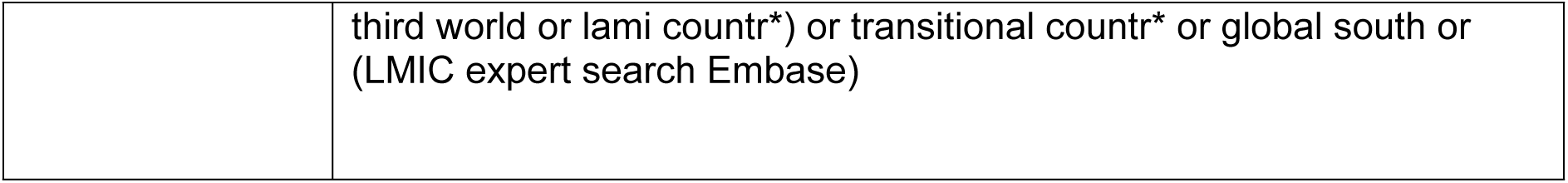
search strategy *Embase*.

**Table 5:** search strategy *Global Health*.

|  |  |
| --- | --- |
| **5. Health service search terms** | (Health service* or health system or health care or healthcare or community health or treatment or therapy or care or mental health or psychotherap* or psychological or psychosocial or reproductive health or "sexual and reproductive" or contraceptiv* or family planning or abortion or post exposure prophylaxis or PEP or support group or hospital* or humanitarian) or health services/ or health care |
|  | AND |
| **6. VAW search terms** | (Gender-based violence or sexual violence or GBV or SGBV or intimate partner violence or rape or "sexual abuse and exploitation" or Domestic violence or family violence or violence against women or VAW or sexual abuse) or domestic violence or intimate partner violence/ or (intimate partner violence or sexual abuse) |
|  | AND |
| **7. Outbreak search terms** | (Outbrea* or epidemi* or pandemic* or public health emergency or Ebola or Filovirus or COVID-19 or Coronavirus or COVID or Zika*) or epidemics/ or Ebolavirus.od. or Ebola haemorrhagic fever/ or outbreaks.sh. or pandemics/ or coronavirus disease 2019/ or Zika virus/ or Zika fever |
|  | AND |
| **8. LMIC search terms** | developing countries/ or least developed countries/ or Threshold Countries/ or ((developing or less* developed or under developed or underdeveloped or middle income or low* income) adj (economy or economies)) or ((developing or less* developed or under developed or underdeveloped or middle income or low* income or underserved or under served or deprived or poor*) adj (countr* or nation? or population? or world)) or (low* adj (gdp or gnp or gross domestic or gross national)) |

**Table 6:** search strategy *Global Index Medicus*.

|  |  |
| --- | --- |
| **1. Health service search terms** | "Health service*" OR "health service* delivery" OR "health system" OR "health care" OR healthcare OR "community health" OR treatment OR therapy OR care OR "mental health" OR psychotherap* OR psychological OR psychosocial OR "reproductive health" OR "sexual and reproductive" OR contraceptiv* OR "family planning" OR abortion OR "post exposure prophylaxis" OR PEP OR "support group" OR hospital* OR humanitarian or MESH terms: "Delivery of Health Care" OR "health services" |
|  | AND |
| **2. VAW search terms** | "Gender-based violence" OR "gender based violence" OR "sexual violence" OR GBV OR SGBV OR "sexual and gender based violence" OR "intimate partner violence" OR rape OR "sexual abuse and exploitation" OR "Domestic violence" OR "family violence" OR "violence against women" OR VAW OR "violence against women and girls" OR "sexual abuse" or MESH terms: "domestic violence" OR "battered woman" OR "partner violence" |
|  | AND |
| **3. Outbreak search terms** | Outbrea* OR epidemi* OR pandemic* OR "public health emergency" OR Ebola OR "Ebola virus disease" OR Filovirus OR COVID-19 OR Coronavirus OR COVID OR Zika* OR MESH terms: epidemic OR Ebola OR "hemorrhagic fever" OR pandemic OR "coronavirus disease" OR "Zika fever" |
|  | AND |
| **4. LMIC search terms** | N/A (database only contains LMIC research) |

